# Cell Catcher: a new method to extract and preserve live renal cells from urine

**DOI:** 10.1101/2023.09.22.23295442

**Authors:** Katia Nazmutdinova, Cheuk Yan Man, Martyn Carter, Philip L Beales, Paul JD Winyard, Stephen B Walsh, Karen L Price, David A Long

**Affiliations:** Developmental Biology and Cancer Research and Teaching Department, University College London Great Ormond Street Institute of Child Health, London, WC1N 1EH, United Kingdom; UCL Centre for Kidney and Bladder Health, London, UK; B-made, The Bartlett School of Architecture, University College London, WC1H 0QB; Genetics and Genomic Medicine Research and Teaching Department, University College London Great Ormond Street Institute of Child Health, London, WC1N 1EH, United Kingdom; London Tubular Centre, Department of Renal Medicine, University College London Medical School, London, UK; Encelo Laboratories Ltd, UK

**Keywords:** human podocytes, human proximal tubule cells, kidney disease, personalised medicine, urine, urine-derived human cells

## Abstract

Patient-specific urine-derived cells are valuable tools for biomedical research and personalised medicine since collection is non-invasive and easily repeated, unlike biopsies. The full potential of urine-derived cells remains untapped, however, due to the short shelf life of samples and necessity for prompt centrifugation. This study aims to address this limitation by evaluating a novel filtration-based Cell Catcher device and comparing its efficiency to centrifugation. We obtained urine from 18 tubulopathy patients and using paired analysis demonstrated that the Cell Catcher device significantly improves the success rate of isolating viable renal cells, and the cell yield. The findings were confirmed in a second independent study, using 44 samples obtained from healthy controls or patients with Bardet-Biedl syndrome or tubulopathies, where colonies were established in 90% of the Cell Catcher-processed samples. Cultured cells displayed a variety of morphologies and expressed markers of podocyte and proximal tubule cells. Collectively, we describe an improved, point-of-care methodology to obtain live patient cells from urine using a filtration technique, with potential personalised medicine applications in nephrology, regenerative medicine, and urological cancers.

## Introduction

A small proportion of urinary tract cells are shed into the urine via normal physiological processes. These include epithelial cells from proximal tubule, Loop of Henle, distal tubule, glomerular podocytes, as well as immune and bladder cells^1^. Urine-derived cells offer advantages over biopsies because they are easily obtained repeatedly without pain or discomfort. Furthermore, they have multiple research uses including “disease in a dish” approaches, to study, model, and screen genetic kidney disorders^2^, generating induced pluripotent stem cells^3^ and as a drug-screening platform^4,5^. Despite these advantages, the full potential of urine-derived cells is not currently being realised.

One key issue is inconsistencies in different methods to initiate cell culture from urine. Urine from healthy adults contains between 2.5-7.5 cells/100 ml, which can proliferate in culture, yielding millions of cells within 2-4 weeks^6,7^. However, the success rate of initiating and expanding cells from urine is variable, ranging from 10-73%, depending on the study and the health status of the individuals^3,8–10^. In addition, the cell population obtained is heterogeneous, containing a mixture of differentiated and undifferentiated renal cells^11^, complicating the interpretation of studies.

These inconsistencies in the reported yields, and the identity of the cell types obtained, are likely due to methodological differences, including culture conditions. Currently, urinary cells are isolated within 4 hours of sample collection, using 2-step centrifugation, which requires a laboratory to be in close proximity to the collection site. In addition to making the process logistically challenging, extended cell exposure to urine in this approach adversely impacts cell viability^1^. The ability to process samples on site quickly could improve yields and standardise protocols, improving reproducibility and facilitating comparisons between studies.

We hypothesised that immediate processing through filtration should improve cell yields by minimising exposure time to urine. To test this, we developed a filtration-based Cell Catcher device for processing urine at clinical sites where samples are collected and directly compared its efficiency to centrifugation.

## Methods

Urine was collected from consented patients at the Royal Free Hospital, St Thomas Hospital and Great Ormond Street Hospital, London (Ethical approval references: 05/Q0508/6, 08/H0713/82).

First, samples from 18 tubulopathy patients (**Table 1**), were split for paired analyses: one processed by the Cell Catcher device (**Fig.1A**) within 30 minutes of collection at the clinic and the other transported on ice to a laboratory to be centrifuged within 4 hours (**Fig.1B**), using 2-step centrifugation at room temperature (10 minutes at 400*g,* and 200*g)* to include a PBS wash, as per published protocols^12^.

**Table 1.** Split sample study: a summary of donor profile, urine sample profile and urine-derived cell culturing outcome. Number of colonies per sample is the average of the counts by two investigators on Day 6 of cell culture. *SG – Specific Gravity, Cont. – contamination, BBS – Bardet-Biedl Syndrome*

| Sample ID | Donor profile |  |  | Urine sample profile |  | Cell culturing outcome |  |
| --- | --- | --- | --- | --- | --- | --- | --- |
|  | Age group | Gender | Clinical diagnosis | Volume, ml | SG | Processed by Cell Catcher, 1/2 of the total volume, (number of colonies) | Processed by Centrifugation, 1/2 of the total volume, (number of colonies) |
| 1 | 50-69 | M | Hypokalaemic Hypertension | 85 | 1.020 | Cont. | Cont. |
| 2 | 30-49 | F | Medullary Nephrocalcinosis | 50 | 1.030 | (0) | (0) |
| 3 | 70-89 | F | Tubulointerstitial Nephritis | 30 | 1.020 | Cont. | Cont. |
| 4 | 50-69 | F | IgA nephropathy | 80 | 1.020 | (21.5) | (8) |
| 5 | 50-69 | M | Tubulointerstitial Nephritis | 30 | 1.025 | (5.5) | (0) |
| 6 | 18-29 | F | Distal renal tubular acidosis (dRTA) | 40 | 1.010 | (35) | (25) |
| 7 | 18-29 | M | Distal renal tubular acidosis (dRTA) | 50 | 1.015 | (26) | (18.5) |
| 8 | 30-49 | M | Kidney stones | 28 | 1.015 | (0) | (0) |
| 9 | 50-69 | F | Distal Salt Losing Tubulopathy | 38 | 1.020 | (0) | (0) |
| 10 | 30-49 | F | Nephrogenic Diabetes Insipidus | 75 | 1.005 | (0) | (0) |
| 11 | 50-69 | F | Tubulointerstitial Nephritis | 20 | 1.010 | (2) | (0) |
| 12 | 30-49 | M | Distal salt losing tubulopathy | 40 | 1.025 | (1) | (0) |
| 13 | 50-69 | F | Tubulointerstitial Nephritis | 80 | 1.025 | (10) | (3) |
| 14 | 30-49 | F | Not yet diagnosed | 100 | 1.015 | (1.5) | (1) |
| 15 | 30-49 | F | Distal salt losing tubulopathy | 100 | 1.015 | Cont. | Cont. |
| 16 | 18-29 | M | Proximal Tubulopathy | 100 | 1.020 | (86) | (60.5) |
| 17 | 50-69 | M | Distal Tubulopathy | 60 | 1.020 | (6.5) | (1) |
| 18 | 18-29 | M | Proximal tubulopathy | 60 | 1.025 | (127.5) | (83) |

**Figure 1.**
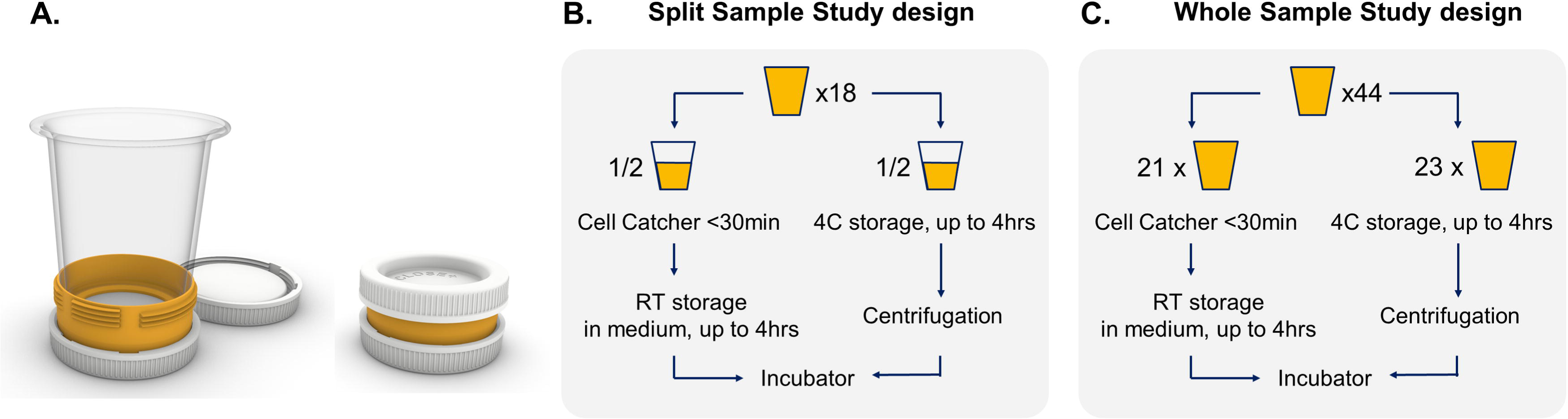
Cell Catcher prototype and studies design. **A.** Cell Catcher diagram. The hub of the Cell Catcher has detachable lids and houses a membrane. It connects to a detachable funnel for urine samples to be processed by gravity. Following filtration, medium is added for the cells to be preserved during transport, inside the hub. Prototypes were produced using Polyjet 3D printing (UCL, B-made 3D printing, Bartlett School of Architecture). **B.** Split sample study design. Eighteen samples were collected from patients with renal tubulopathies, each sample was split into two equal parts, Cell Catcher group and centrifugation group. Samples fractions in the Cell Catcher group were processed on site within 30 minutes of collection and stored at room temperature for up to 4 hours during transportation to the laboratory, where they were plated. Samples fractions in the centrifugation group were stored at 4C and transported to the laboratory on ice within 4 hours to be centrifuged and plated. **C.** Whole sample study design. 44 samples were collected from patients with renal tubulopathies, BBS and healthy adults and whole volume was processed either by a Cell Catcher or centrifugation.

In a second study, we assessed samples from tubulopathy patients (*n=18*), adult and paediatric patients with Bardet-Biedl Syndrome (*n=15*) and healthy controls (*n=11*) (**Table 2**). In these 44 individuals, the whole sample volume were processed either by Cell Catcher or centrifugation (**Fig.1C**).

**Table 2.**
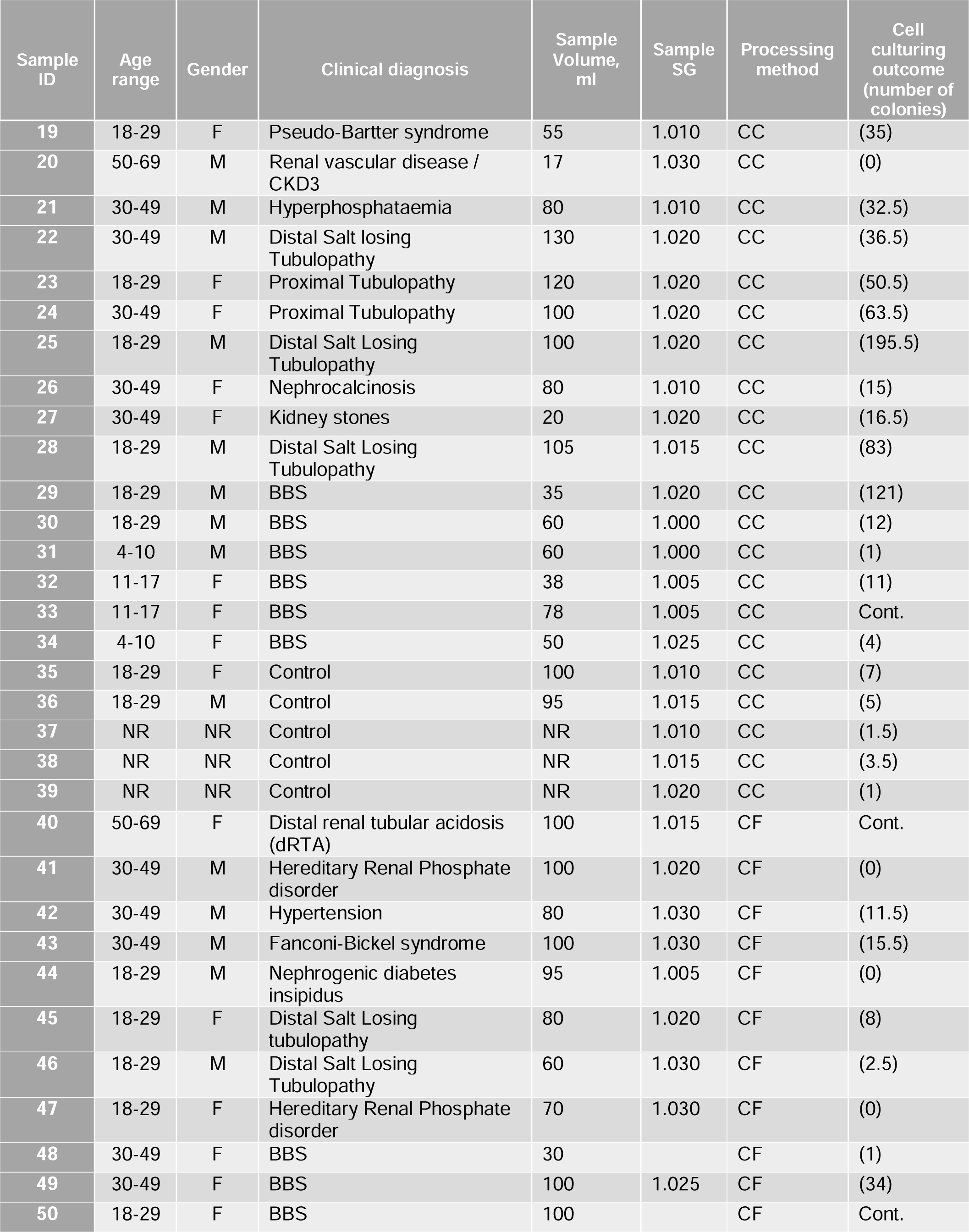
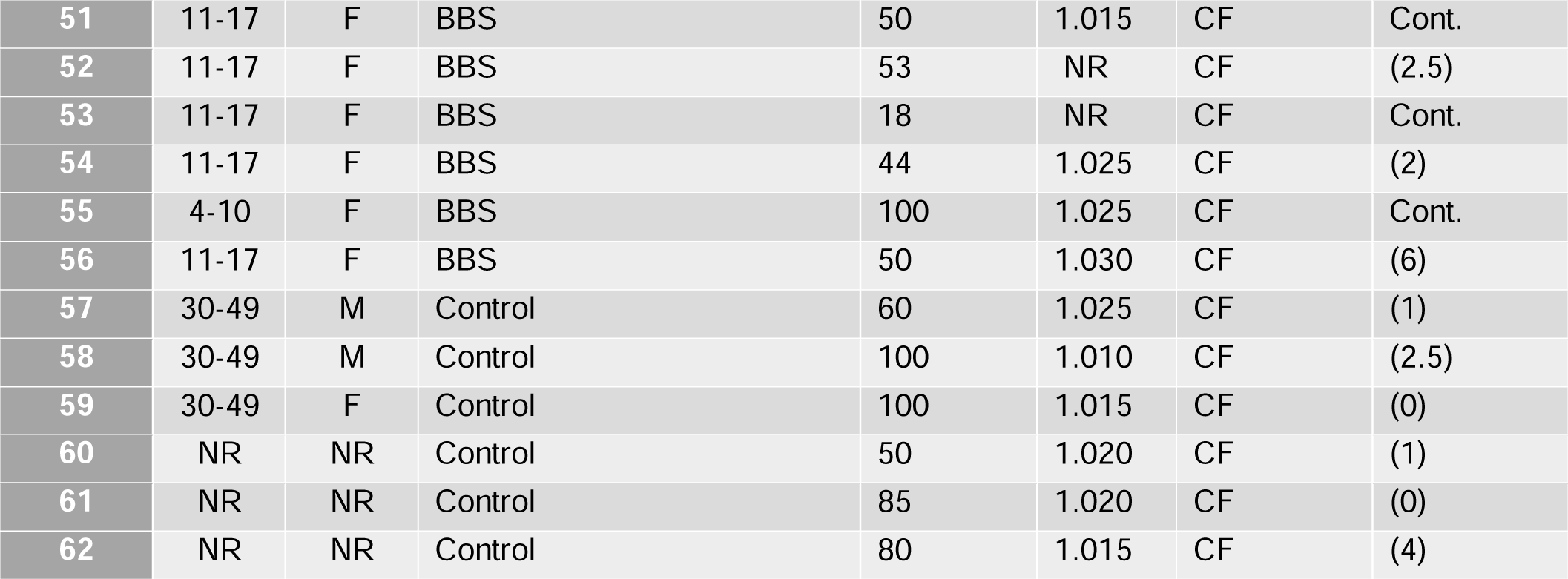
Whole sample study: a summary of donor profile, urine sample profile and urine-derived cell culturing outcome. Number of colonies per sample is the average of the counts by two investigators on Day 6 of cell culture*. SG – Specific Gravity, CC-Cell Catcher. CF – Centrifugation, BBS – Bardet-Biedl Syndrome, CKD – Chronic Kidney Disease, Cont. - contamination, NR – not recorded*

| Sample ID | Age range | Gender | Clinical diagnosis | Sample Volume, ml | Sample SG | Processing method | Cell culturing outcome (number of colonies) |
| --- | --- | --- | --- | --- | --- | --- | --- |
| 19 | 18-29 | F | Pseudo-Bartter syndrome | 55 | 1.010 | CC | (35) |
| 20 | 50-69 | M | Renal vascular disease / CKD3 | 17 | 1.030 | CC | (0) |
| 21 | 30-49 | M | Hyperphosphataemia | 80 | 1.010 | CC | (32.5) |
| 22 | 30-49 | M | Distal Salt losing Tubulopathy | 130 | 1.020 | CC | (36.5) |
| 23 | 18-29 | F | Proximal Tubulopathy | 120 | 1.020 | CC | (50.5) |
| 24 | 30-49 | F | Proximal Tubulopathy | 100 | 1.020 | CC | (63.5) |
| 25 | 18-29 | M | Distal Salt Losing Tubulopathy | 100 | 1.020 | CC | (195.5) |
| 26 | 30-49 | F | Nephrocalcinosis | 80 | 1.010 | CC | (15) |
| 27 | 30-49 | F | Kidney stones | 20 | 1.020 | CC | (16.5) |
| 28 | 18-29 | M | Distal Salt Losing Tubulopathy | 105 | 1.015 | CC | (83) |
| 29 | 18-29 | M | BBS | 35 | 1.020 | CC | (121) |
| 30 | 18-29 | M | BBS | 60 | 1.000 | CC | (12) |
| 31 | 4-10 | M | BBS | 60 | 1.000 | CC | (1) |
| 32 | 11-17 | F | BBS | 38 | 1.005 | CC | (11) |
| 33 | 11-17 | F | BBS | 78 | 1.005 | CC | Cont. |
| 34 | 4-10 | F | BBS | 50 | 1.025 | CC | (4) |
| 35 | 18-29 | F | Control | 100 | 1.010 | CC | (7) |
| 36 | 18-29 | M | Control | 95 | 1.015 | CC | (5) |
| 37 | NR | NR | Control | NR | 1.010 | CC | (1.5) |
| 38 | NR | NR | Control | NR | 1.015 | CC | (3.5) |
| 39 | NR | NR | Control | NR | 1.020 | CC | (1) |
| 40 | 50-69 | F | Distal renal tubular acidosis (dRTA) | 100 | 1.015 | CF | Cont. |
| 41 | 30-49 | M | Hereditary Renal Phosphate disorder | 100 | 1.020 | CF | (0) |
| 42 | 30-49 | M | Hypertension | 80 | 1.030 | CF | (11.5) |
| 43 | 30-49 | M | Fanconi-Bickel syndrome | 100 | 1.030 | CF | (15.5) |
| 44 | 18-29 | M | Nephrogenic diabetes insipidus | 95 | 1.005 | CF | (0) |
| 45 | 18-29 | F | Distal Salt Losing tubulopathy | 80 | 1.020 | CF | (8) |
| 46 | 18-29 | M | Distal Salt Losing Tubulopathy | 60 | 1.030 | CF | (2.5) |
| 47 | 18-29 | F | Hereditary Renal Phosphate disorder | 70 | 1.030 | CF | (0) |
| 48 | 30-49 | F | BBS | 30 |  | CF | (1) |
| 49 | 30-49 | F | BBS | 100 | 1.025 | CF | (34) |
| 50 | 18-29 | F | BBS | 100 |  | CF | Cont. |
| 51 | 11-17 | F | BBS | 50 | 1.015 | CF | Cont. |
| 52 | 11-17 | F | BBS | 53 | NR | CF | (2.5) |
| 53 | 11-17 | F | BBS | 18 | NR | CF | Cont. |
| 54 | 11-17 | F | BBS | 44 | 1.025 | CF | (2) |
| 55 | 4-10 | F | BBS | 100 | 1.025 | CF | Cont. |
| 56 | 11-17 | F | BBS | 50 | 1.030 | CF | (6) |
| 57 | 30-49 | M | Control | 60 | 1.025 | CF | (1) |
| 58 | 30-49 | M | Control | 100 | 1.010 | CF | (2.5) |
| 59 | 30-49 | F | Control | 100 | 1.015 | CF | (0) |
| 60 | NR | NR | Control | 50 | 1.020 | CF | (1) |
| 61 | NR | NR | Control | 85 | 1.020 | CF | (0) |
| 62 | NR | NR | Control | 80 | 1.015 | CF | (4) |

Cell Catcher is a patent-pending (Encelo Laboratories Ltd), custom-built device manufactured using 3D printing that houses a polyethersulfone 5µm membrane (Sterlitech, Auburn, USA). Gravity-fed filtration is achieved for samples of up to 100ml in volume with low-specific gravity (SG, 1.005-1.015), or for smaller volumes (<25ml) of high-SG samples (1.020-1.030).

Standard Encelo medium was used to resuspend cells following filtration/centrifugation, and to culture them. It consists of DMEM High Glucose/F12 (1:1), 1% Penicillin/Streptomycin, 1% Amphotericin B, 10% FBS with added growth factors (human epidermal growth factor, insulin, hydrocortisone, transferrin, triiodothyronine, epinephrine, bovine pituitary extract and adenine). Cell suspensions were cultured at 37°C in a 5% CO_2_ incubator. Half of the media was replaced with fresh one daily until day 3 and replaced completely every 2 days thereafter. Cells were passaged at around 80% confluence.

Cultures were monitored daily and assigned to the following categories: contaminated, formed colonies, no colonies. Where colonies (cell cluster of >10cells) have formed, the number was counted on day 6 of culture independently by two investigators. RNA was extracted at first passage of the cells using the RNeasy Plus Mini kit (Qiagen, Hilden, Germany); 500ng was then used to synthesise cDNA using the iScript™ gDNA Clear cDNA Synthesis Kit (BioRad, Hercules, USA). Transcripts of established markers of renal and bladder cells^13^ (Wilms tumour 1 (*WT1*), podocin (*NPHS2*), uroplakin 3A (*UPK3A*), uromodulin (*UMOD*), aquaporin-3 (*AQP3*) and aminopeptidase-A (*ENPEP*)) were assessed (**Table 3**) using RT-PCR. SYBR™ Green PCR Master Mix (Thermo Fisher, Waltham USA). The house-keeping gene, glyceraldehyde-3-phosphate dehydrogenase (*GAPDH*) was used as a loading control, with RNA extracted from total kidney and bladder as positive controls.

**Table 3.**
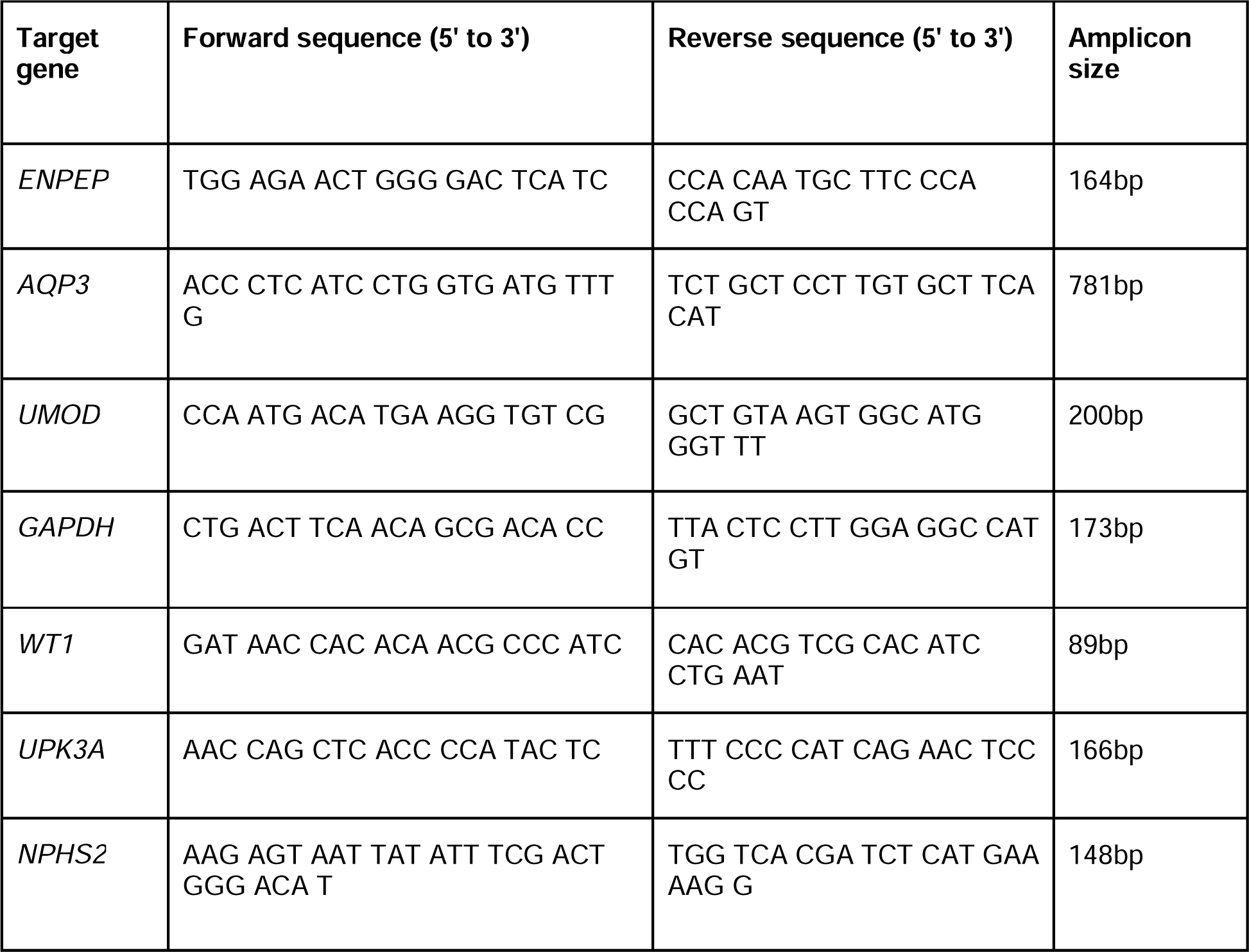
Primer sequences. Wilms tumour 1 (WT1), podocin (NPHS2), uroplakin 3A (UPK3A), uromodulin (UMOD), aquaporin-3 (AQP3) and aminopeptidase-A (ENPEP)

Data normality was assessed using the Shapiro-Wilk test and significance assessed by either *t*-tests or Wilcoxon signed-rank non-parametric test. Statistical significance was accepted at *p*>0.05.

## Results

In a split sample paired comparison 11 (Cell Catcher) versus 8 (centrifugation) samples formed colonies by day 6 (61% versus 44%, respectively), 4 versus 7 did not (22% versus 39%, respectively) and 3 samples were contaminated in each group (**Fig.2A**). In the 11 cultures that formed colonies in the Cell Catcher fraction, we counted the number and compared this directly in a paired analysis with the corresponding centrifugation samples from the same patient. In all eleven patients, the Cell Catcher-processed fraction contained a significantly (p=0.001) greater number of colonies (**Fig.2B**). In 8 samples where colonies have formed in both fractions, the number was on average double in the Cell Catcher-processed fraction (96% increase).

**Figure 2.**
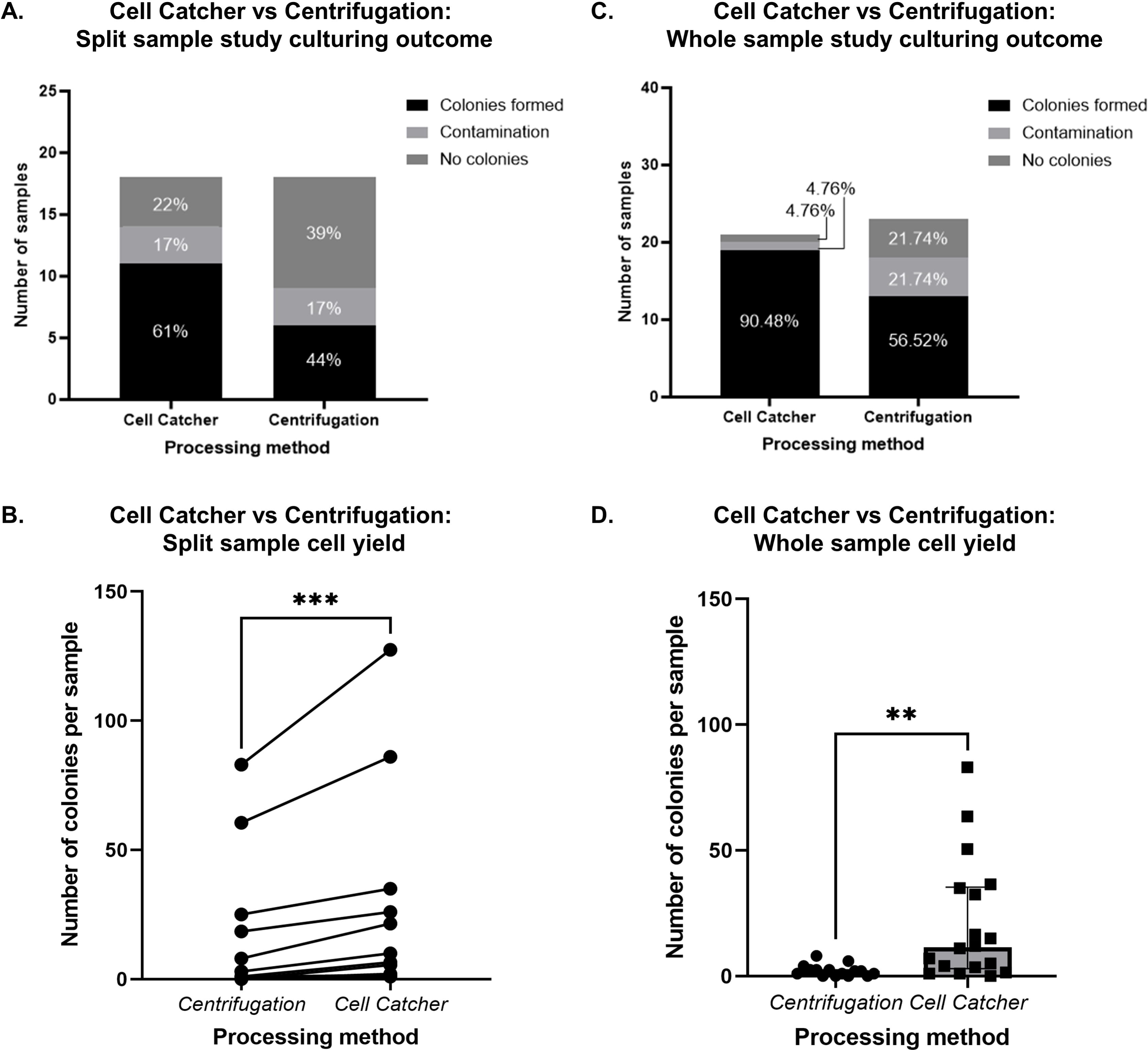
Cell Catcher clinical validation study results. **A.** Culturing outcomes, split sample study. Cells from sample fractions processed by either Cell Catcher or centrifugation were seeded, cultured and assigned to the following categories: no colonies (by 2 weeks), colonies (day 6) and contamination (usually within the first 3 days). Distribution of the three culturing outcomes for each experimental condition is shown. **B.** Split sample cell yield differences between Cell Catcher and centrifugation fractions. Colonies (cell clusters of >10 cells) were quantified on Day 6 after plating, by two researchers independently. Average numbers of the two counts are plotted for each of the 11 samples in the paired study, where colonies formed in at least one of the fractions (Wilcoxon non-parametric paired t-test, n=11, p-value=0.001). **C.** Culturing outcomes, whole sample study. Cells from samples processed by either Cell Catcher or centrifugation were seeded, cultured and assigned to the following categories: no colonies (by 2 weeks), colonies (day 6) and contamination (usually within the first 3 days). Distribution of the three culturing outcomes for each experimental condition is shown. **D.** Whole sample cell yield differences between Cell Catcher and centrifugation-processed samples. Colony counts in samples processed by either Cell Catcher (n=18) versus centrifugation (n=16), after 2 outliers that were identified in each group were removed. Mann Whitney test, p-value 0.0013. Median + interquartile range is plotted.

Similar observations were found in the 44 donors where the whole sample was processed using either Cell Catcher or centrifugation (**Fig.2C**). Here, 90% of Cell Catcher-processed samples (n=21) contained colonies, while colonies were only found in 57% of the 23 samples processed by centrifugation. On average, the Cell-Catcher-processed samples contained significantly higher number of colonies (11.5 versus 2, respectively (p-value=0.0012, **Fig 2D**). There were no significant differences in sample volume, sex ratio, age, clinical diagnoses, but the SG in the Cell Catcher group was lower (1.014 vs 1.021).

Cells isolated using Cell Catcher were successfully expanded with yields of 0.5-2.2 million cells within 2 weeks. Examination of the cells by light microscopy revealed several morphologies (**Fig.3A**). RNA was extracted from seven tubulopathy patient samples and transcripts of nephron segment markers assessed by RT-PCR (**Fig.3B**). In all samples, we detected *WT1* and *ENPEP* suggestive of podocyte and proximal tubule cells. Some samples were also positive for *NPHS2*, but not *AQP3*, *UMOD* or *UPK3A.* Cell cultures became phenotypically homogeneous with time, before ceasing to proliferate by week 4-6.

**Figure 3.**
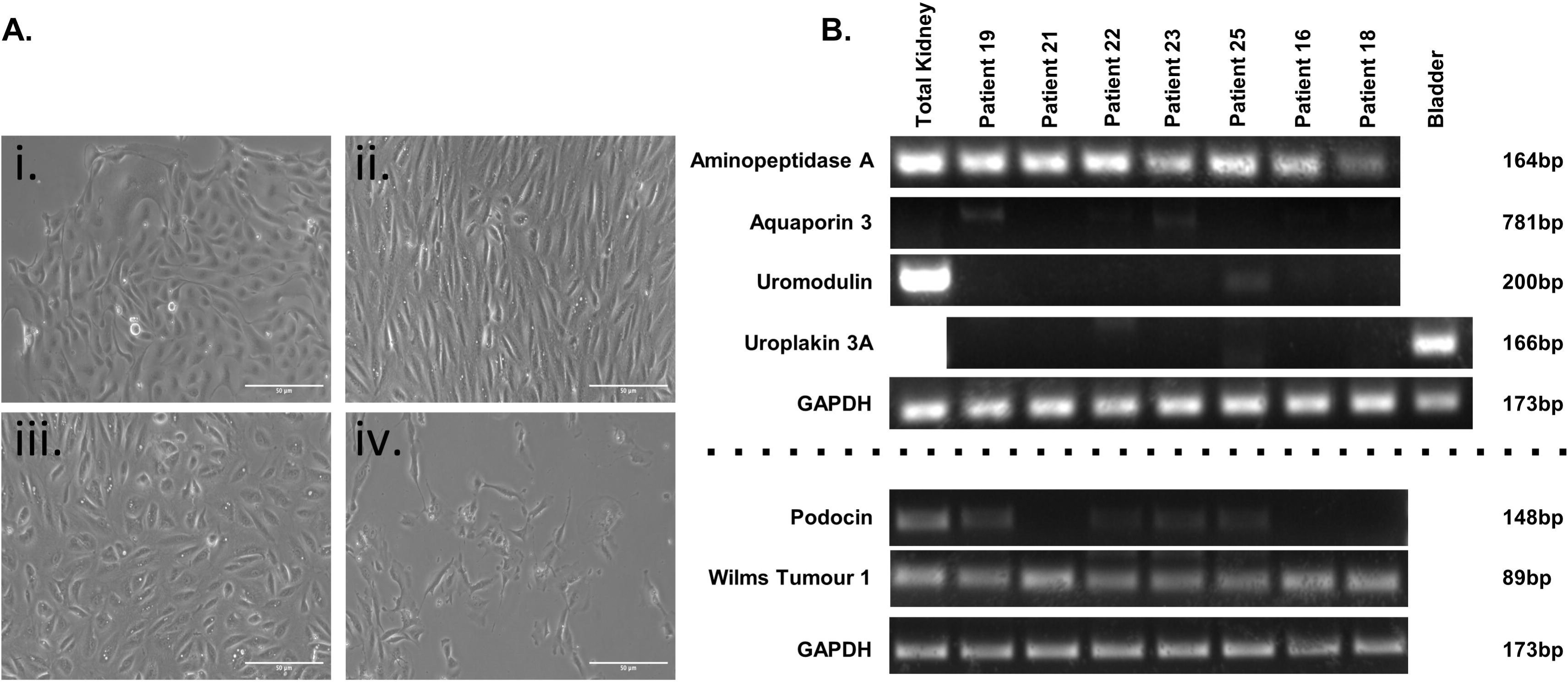
Urine-derived cell characterisation study results. **A.** Representative images of different cell morphologies observed in urine derived cells. Variation in cell and colony morphology was observed, with multiple types sometime present within one sample. Most commonly encountered proliferating types are (i-iii) with (iv) never reaching confluency. Cells with rice-grain morphology (iii) exhibited the best proliferative capacity, eventually becoming elongated (ii) and uniform with repeat passaging. Scale bar = 50µm. **B.** RT-PCR results summary. Tubulopathy patient-derived urinary cells were characterised using RT-PCR to detect transcripts of *ENPEP* (Aminopeptidase A), *AQP3* (Aquaporin 3), *UMOD* (Uromodulin), *UPK3A* (Uroplakin 3A), *NPHS2* (Podocin) and *WT1* (Wilms Tumour 1). *ENPEP* and *WT1* transcripts were detected in all patient samples.

## Discussion

We present a novel Cell Catcher device designed to standardise urine processing methods, reduce the need for urgent laboratory processing and increase cell yield. Where urine samples were split, we report a 17% increase in samples with viable cells using Cell Catcher rather than centrifugation, with a two-fold increase of cells capable of attachment and proliferation. The Cell Catcher achieved a 90% colony formation success rate (19/21) when processing whole-volume, lower-SG samples. Overall contamination rate was 14.5% (9/62) and seen predominantly in females (8/9), especially in those with BBS. By defining optimal physical and biochemical urine properties (e.g. SG, volume) and improving collection protocols for easy mid-stream sampling for all, a higher success rate could potentially be realised. Our findings indicate that immediate on-site processing by filtration is an easier, quicker and more efficient method to concentrate cells from urine samples.

Urine-derived cells are well suited to study inherited kidney disease, as remarkably high number of physiologically relevant cells is detected in patients with or being predisposed to renal dysfunctions caused by genetic conditions (BBS, Bartter Syndrome, Dent’s Disease). While up to 12 colonies can be established from healthy people, some patients shed more than 200 colony-initiating cells in one 100ml sample. Similarly, higher yields have been observed in patients with other renal conditions (nephrotic syndrome, kidney stones, and Fanconi syndrome^8,10,14,15^). We detected transcripts of proximal tubule cell markers in all urine samples, in addition to podocyte markers in some, confirming heterogeneity of cell populations shed in urine. Further research is needed to explore the device’s compatibility with other cell types and downstream processing workflows, including refinement of culturing conditions and medium formulations to isolate and/or expand other cell types known to be present in urine. In addition, the current Cell Catcher version’s functionality is dependent on a sample’s SG: it works best with low-SG samples which usually correlates with a high hydration level of the donor.

The Cell Catcher offers a distinct advantage over centrifugation by its potential compatibility with home use. By introducing the right preservation medium, the time frame for processing filtered samples can be extended, enabling direct shipment of live cells by patients. This innovation has the potential to advance methods of primary cell acquisition, by offering non-invasive, remote and scalable procurement of live cells for clinical and research applications in the renal field and beyond.

## Data Availability

All data produced in the present work are contained in the manuscript.

## Acknowledgements

This work was supported by Encelo Laboratories Ltd, a Kidney Research UK Innovation Grant (Paed_IN_005_20190926 to DAL, PB and JN) and the NIHR Biomedical Research Centre at Great Ormond Street Hospital for Children NHS Foundation Trust and University College London.

## References

1. Abedini A, Zhu YO, Chatterjee S, et al. Urinary Single-Cell Profiling Captures the Cellular Diversity of the Kidney. Journal of the American Society of Nephrology. 2021;32(3):614–627. doi:10.1681/ASN.2020050757

2. Bondue T, Arcolino FO, Veys KRP, et al. Urine-Derived Epithelial Cells as Models for Genetic Kidney Diseases. Cells. 2021;10(6):1413. doi:10.3390/cells10061413

3. Zhou T, Benda C, Dunzinger S, et al. Generation of human induced pluripotent stem cells from urine samples. Nat Protoc. 2012;7(12):2080–2089. doi:10.1038/nprot.2012.115

4. Schutgens F, Rookmaaker MB, Margaritis T, et al. Tubuloids derived from human adult kidney and urine for personalized disease modeling. Nat Biotechnol. 2019;37(3):303–313. doi:10.1038/s41587-019-0048-8

5. Falzarano MS, D’Amario D, Siracusano A, et al. Duchenne Muscular Dystrophy Myogenic Cells from Urine-Derived Stem Cells Recapitulate the Dystrophin Genotype and Phenotype. Hum Gene Ther. 2016;27(10):772–783. doi:10.1089/hum.2016.079

6. Zhang Y, McNeill E, Tian H, et al. Urine Derived Cells are a Potential Source for Urological Tissue Reconstruction. Journal of Urology. 2008;180(5):2226–2233. doi:10.1016/j.juro.2008.07.023

7. Wu S, Liu Y, Bharadwaj S, Atala A, Zhang Y. Human urine-derived stem cells seeded in a modified 3D porous small intestinal submucosa scaffold for urethral tissue engineering. Biomaterials. 2011;32(5):1317–1326. doi:10.1016/j.biomaterials.2010.10.006

8. Wilmer MJG, De Graaf-Hess A, Blom HJ, et al. Elevated oxidized glutathione in cystinotic proximal tubular epithelial cells. Biochem Biophys Res Commun. 2005;337(2):610–614. doi:10.1016/j.bbrc.2005.09.094

9. Wilmer MJ, Saleem MA, Masereeuw R, et al. Novel conditionally immortalized human proximal tubule cell line expressing functional influx and efflux transporters. Cell Tissue Res. 2010;339(2):449–457. doi:10.1007/s00441-009-0882-y

10. Dörrenhaus A, Müller JIF, Golka K, Jedrusik P, Schulze H, Föllmann W. Cultures of exfoliated epithelial cells from different locations of the human urinary tract and the renal tubular system. Arch Toxicol. 2000;74(10):618–626. doi:10.1007/s002040000173

11. Oliveira Arcolino F, Tort Piella A, Papadimitriou E, et al. Human urine as a noninvasive source of kidney cells. Stem Cells Int. 2015;2015. doi:10.1155/2015/362562

12. Guo Z, Su W, Zhou R, et al. Exosomal MATN3 of Urine-Derived Stem Cells Ameliorates Intervertebral Disc Degeneration by Antisenescence Effects and Promotes NPC Proliferation and ECM Synthesis by Activating TGF-β. Oxid Med Cell Longev. 2021;2021:1–18. doi:10.1155/2021/5542241

13. Price KL, Kolatsi-Joannou M, Mari C, Long DA, Winyard PJD. Lithium induces mesenchymal-epithelial differentiation during human kidney development by activation of the Wnt signalling system. Cell Death Discov. 2018;4(1):13. doi:10.1038/s41420-017-0021-6

14. Lazzeri E, Ronconi E, Angelotti ML, et al. Human Urine-Derived Renal Progenitors for Personalized Modeling of Genetic Kidney Disorders. J Am Soc Nephrol. Published online 2015:1–14. doi:10.1681/ASN.2014010057

15. Inoue CN, Sunagawa N, Morimoto T, et al. Reconstruction of tubular structures in three-dimensional collagen gel culture using proximal tubular epithelial cells voided in human urine. In Vitro Cell Dev Biol Anim. 2003;39(8-9):364–367. doi:10.1290/1543-706X(2003)0392.0.CO;2

